# JYNNEOS vaccination induced short-lived neutralizing antibody responses to monkeypox virus in naïve individuals

**DOI:** 10.1101/2024.08.28.24312561

**Authors:** Kara Phipps, Jennifer Yates, Jessica Machowski, Sean Bialosuknia, Danielle Hunt, Alan DuPuis, Anne Payne, William Lee, Kathleen A. McDonough

**Author notes:** Corresponding author: Kathleen A. McDonough, PhD, Wadsworth Center, NYSDOH, 120 New Scotland Ave, Albany NY 12208. **Author Contributions:** KP contributed to the experimental design, data acquisition and analysis, interpretation of data and writing of manuscript; JY contributed to conception of work, experimental design, interpretation of data and writing of manuscript; JM, SB, and DH contributed to data acquisition and analysis; AD, AP, WL, contributed to conception of work, experimental design, and interpretation of data. KM contributed to conception of work, experimental design, and interpretation of data, and writing of the manuscript.

## Abstract

Current studies of the JYNNEOS-induced neutralizing antibody response to monkeypox virus (MPXV) are limited by either short-term durability data, quantification in an endemic population, or lack of an infectious MPXV neutralization assay. We used plaque reduction neutralization test (PRNT) with authentic MPXV and vaccinia viruse (VACV) to assess antibody responses over twelve months of eight donors vaccinated with two doses of JYNNEOS. One donor previously received the ACAM2000 vaccine; seven donors were smallpox-vaccine naïve. The IgG response of the donors to VACV (L1R, B5R, and A33R) and MPXV (E8L, H3L, A35R) antigens and PRNT titers to both viruses peaked at eight weeks post-vaccination and waned thereafter in naïve donors. MPXV PRNT titers were especially low; no naïve donors produced a detectable PRNT90 titer. Our results suggest the MPXV humoral response produced by JYNNEOS is limited in naïve donors and invites further investigation into current mpox vaccination strategies and correlates of protection.

## Introduction

Monkeypox virus (MPXV), the causative agent of mpox disease, is an emerging public health concern. MPXV is considered endemic in areas of central and western Africa, where it causes thousands of cases annually (1). In 2022, a large global outbreak of mpox spread primarily among men who have sex with men. The 2022 outbreak heightened awareness of the need for preventative measures against transmission and severe mpox disease, triggering a public health campaign that included recommending behavioral changes and vaccination with the Modified vaccinia Ankara-Bavarian Nordic (MVA-BN) vaccine for populations most at risk. Little was known about the efficacy or durability of the neutralizing antibody response produced by MVA-BN against MPXV at the time of this global outbreak.

MPXV is a member of the *Orthopoxvirus* genus and is related to variola virus, the causative agent of smallpox, as well as less virulent genus members including cowpox and vaccinia (VACV). Due to their relatedness, immunity elicited from vaccinia-based smallpox vaccines produces cross reactivity against MPXV (2, 3). First and second generation smallpox vaccines comprised of replication competent strains of vaccinia are currently not recommended to the general population due to potentially severe or fatal side effects for some individuals, including those infected with HIV (4).

MVA-BN is considered a safer, “third generation” smallpox vaccine as it is a highly passaged vaccinia strain that does not replicate in humans. Due to the public health urgency, MVA-BN was approved for vaccination to protect against mpox disease in the United States and Europe prior to fully establishing the potency and durability of its neutralizing antibody response to MPXV in humans of a non-endemic region (5, 6). The United States Food and Drug Administration (FDA) approved use of MVA-BN under the name JYNNEOS in 2019, while the European Medicines Agency (EMA) approved it under the name IMVANEX in 2022.

Epidemiological studies from the US support vaccine efficacy for MVA-BN and have estimated its effectiveness against MPXV to range from 66 to 88.5% in fully vaccinated individuals (7-10). However, the majority of mpox cases captured occurred less than six months after the peak of MVA-BN vaccine administration in the US, and the potential for waning efficacy was not assessed in these studies. Impacts of behavioral changes are also hard to quantify and the role of MVA-BN in quelling the mpox outbreak in the US has been challenged by modeling of infection rates during the 2022 epidemic (11). Paredes et al [11] concluded that mpox transmission dropped dramatically before vaccination-induced immunity could play a role. Multiple cases of breakthrough mpox infection have been reported to occur within a year of MV-BN vaccination response, raising further questions about the efficacy and durability of the vaccine-induced immune response (12). Here we characterize the efficacy and durability of neutralizing antibody (nAb) responses generated by the JYNNEOS vaccine to MPXV in a small cohort of donors using a native MPXV plaque reduction neutralization test (PRNT). We found that vaccinees without known exposure to earlier smallpox vaccines had limited neutralizing capacity against MPXV which waned in less than a year.

## Materials and Methods

### Human Subjects

This assay development study was performed using deidentified sera and plasma for a public health function in a declared Public Health Emergency (PHE). It has been deemed “Non-human subject research” by the NYS Institutional Review Board. The vaccinee cohort is composed of 8 serum specimens from NYS Department of Health employee donors who were vaccinated with JYNNEOS® because of potential occupational exposure.

### Viruses and Cells

The following reagents were obtained through BEI Resources, NIAID, NIH: Vaccinia Virus, Western Reserve (NIAID, Tissue Culture Adapted), NR-55 and Monkeypox Virus, USA-2003, NR-2500. Virus stocks were passaged once in Vero E6 cells (African green monkey kidney, ATCC CRL-1587) maintained in Eagle’s Minimum Essential Medium (EMEM) with 2% heat-inactivated fetal bovine serum, Penicillin (100 unit/mL) and Streptomycin (100 μg/mL).

### Sonication

Sonication was performed in sealed tubes with the Virtis Virsonic 100 cup horn sonicator continuously cooled to 4°C with a circulating water bath. Virus was diluted in Eagle’s EMEM with 2% fetal bovine serum and separate aliquots were sonicated with increasing intensity at settings 2, 3, 4, and 5 for four five second bursts separated by five second rest intervals to determine optimal sonication conditions.

### Plaque reduction neutralization test (PRNT)

Test serum was not heat inactivated unless otherwise noted. Each serum sample was serially diluted 2-fold in EMEM + 2% FBS. An equal volume of media containing either VACV or MPXV was sonicated at setting 3 and added to each sample at a concentration expected to yield approximately 100 plaque forming units (PFU). The virus:serum mixture was incubated at 37°C for 1 hour, with the exception of experiments in Figure S3, which took place for 24 hours. The mixture was then inoculated onto VeroE6 cell monolayers and adsorbed for 1 hour at 37°C. EMEM media containing 0.6% oxoid agarose was then added to wells, allowed to solidify, and incubated at 37°C with 5% CO_2_. A secondary overlay containing 0.2% neutral red (manufacturer details) was added for plaque visualization at 48 hours post infection. Plaques were counted 24 hours later. Neutralization titers were determined as the serum dilution resulting in a 50% (PRNT_50_) or 90% (PRNT_90_) plaque reduction compared to the virus working dilution (approximately 100-250 PFU). Virus inoculum used to enumerate the working dilution was incubated in media alone alongside virus:sera samples prior to infection and then titrated by plaque assay in parallel to PRNT. Positive and negative control antibodies were included in each assay and a four-fold difference in the range of control antibodies would result in rejection of assay results. PRNT titers measuring the efficacy of JYNNEOS in vaccinated donors over time are the result of two independent experiments, except Figure S3, which represents only a single assay.

### Recombinant OPV Antigens

Recombinant proteins were obtained from several sources. Recombinant A33R (VAC-WR-A33R; Cat# NR545), B5R (VAC-WR-B5R; Cat# NR-546) and L1R (VAC-WR-L2R; Cat# NR-21986) were obtained from BEI Resources, Manassas VA. Mpox A35R (Cat # 230-30238), E8L (Cat# 230-30232) and H3L (Cat# 230-30233) were purchased from Ray Biotech, Peachtree Corners, GA.

### Orthopoxvirus-specific Multiplex Microsphere Immunoassay (MIA)

Specimens were assessed for the presence of antibodies reactive to OPV antigens using an MIA as previously described (12). Recombinant proteins were covalently linked to the surface of fluorescent, magnetic microspheres (Luminex Corporation). Serum or plasma samples (25 μl at 1:100 dilution) and antigen-coupled microspheres (25 μl at 5×10^4^ microspheres/mL per manufacturer instructions) were mixed and incubated for 30 minutes at 37°C. Serum-bound microspheres were washed and incubated with phycoerythrin (PE)-conjugated secondary antibody specific for human IgG (Southern Biotech). After washing and final resuspension in buffer, the samples were analyzed on a FlexMap 3D analyzer using xPONENT software, version 4.3 (Luminex Corporation).

### Calculation of Cutoffs and Index Values

ROC curves were generated in GraphPad Prism 9.1.0 for each antigen based on the MFI values of the 120 mpox-negative donors born after 1970 and 40 mpox-positive confirmed donors. Sensitivity and specificity values generated by the ROC curve were used to calculate cutoffs using a Youden’s J index (J = sensitivity + specificity – 1) for the range of MFI values in the ROC analysis. The cutoff value was set as the MFI equaling the highest Youden’s J index which represents the best balance of specificity and sensitivity over the range of the assay. MFI signals for antigen comparisons were normalized for background fluorescence using an index value (MFI / clinical cutoff).

### Statistical Analyses

One-way ANOVA was used to assess statistical significance. For multiple comparisons of the differences in means of 3 or more groups to a control group, one-way ANOVA was followed by the Dunnett’s multiple comparison test.

## Results

Studies were performed with donated sera from individuals (n=8) immunized with a two-dose regimen of the JYNNEOS vaccine against potential occupational exposure. Vaccine doses were administered approximately 28 days apart and sera were collected from all participants shortly before JYNNEOS vaccination (pre-vax) and at sequential time points until 12 months post-vaccination. Seven donors were administered the vaccine subcutaneously and one donor received the vaccine intradermally. One donor had received ACAM2000, a second-generation smallpox vaccine approximately five years prior to JYNNEOS vaccination. The remaining donors were determined to be previously smallpox-vaccine naïve by a combination of personal account, lack of a vaccine “take” scar, and/or age. Due to differences in timing of immunization, the 12-month sampling point included sera for only seven of the eight participants.

Donor sera were examined for IgG reactivity to MPXV- and VACV -derived antigens by using a previously described microsphere immunoassay (13) to assess overall antibody levels and cross-reactivity to MPXV in response to JYNNEOS vaccination. Orthopoxvirus virions have two forms, which differ in their surface proteins: intracellular mature virions (IMV) and extracellular enveloped virions (EEV), so antigens from each form were tested. VACV L1R and MPXV E8L are found on IMVs, while the remaining antigens are found on EEVs. VACV recombinant proteins L1R, A33R, and B5R were selected for quantification as immunization by these antigens and VACV A27L demonstrated protection from lethal mpox in nonhuman primates (14). MPXV recombinant protein antigens were chosen by availability.

Sera from the donor with prior smallpox vaccination (Fig 1A-B) displayed much higher IgG reactivity than the naïve donors and was excluded from the mean values in Fig 1C-D. In naïve donors, the mean sera IgG reactivity became positive for all VACV antigens tested with VACV L1R showing the highest mean IgG reactivity of all antigens tested (Fig 1D). In contrast, E8L was the sole MPXV antigen with positive mean IgG reactivity in naïve donors despite MPXV A35R being homologous to VACV A33R (Fig 1C). For all antigens, IgG reactivity peaked at approximately eight weeks post initial dose and waned thereafter, indicating the antibody response generated by JYNNEOS is short-lived in naive individuals (15). In contrast, sera from the previously vaccinated individual remained stably positive beyond 250 days post-vaccination for all antigens with the exception of MPXV A35R (Fig 1A-B). We also noted that sera from naive individuals reacted most strongly to the IMV antigens from both viruses (L1R and E8L). This bias towards MPXV IMV antigen E8L was not present in sera from the previously vaccinated individual despite their response to VACV L1R being more robust.

**Figure 1:**
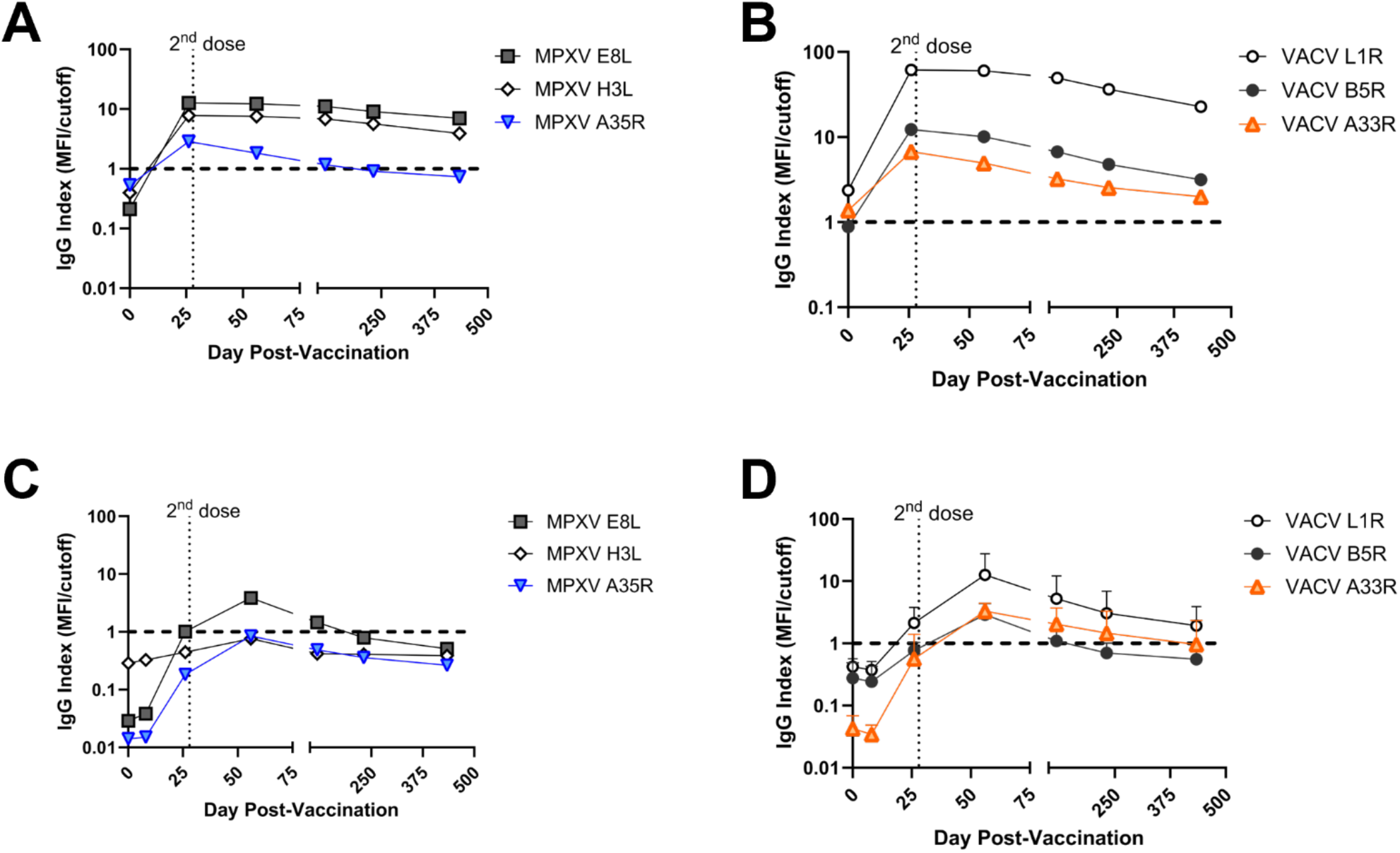
IgG antibody reactivity to orthopoxvirus antigens in JYNNEOS vaccinees with and without prior smallpox vaccination. Serum specimens from eight JYNNEOS vaccine recipients were analyzed for IgG reactivity to recombinant protein antigens derived from mpox or vaccinia virus by microsphere immunoassay. One donor with who received ACAM2000 prior to JYNNEOS is shown separately in A and B. Means of seven individuals who had no prior smallpox vaccination are shown in C and D. Mean index values (MFI/cutoff) of MPXV E8L (gray squares), MPXV A35R (blue triangles) and MPXV H3L (white diamonds) were plotted for days 0, 8, 26, 56, 118, 231, and 434 post-vaccination (A,C). Mean index values (MFI/cutoff) of VACV L1R (white circles), VACV A33R (orange triangles) and VACV B5R (gray circles) were plotted for 0, 8, 26, 56, 118, 231, and 434 post-vaccination (B, D). The black dashed line at y = 1.0 indicates the cutoff value. The dotted line indicates the second dose of vaccine at day 28 post vaccination.

The PRNT is considered the gold standard for measuring neutralizing antibody levels as it directly measures inhibition of native virus infection. Our PRNT was developed by minor modifications of a standard assay (16, 17).

Orthopoxviruses such as VACV are known to form multi-virion aggregates (18, 19) and such structures are capable of affecting antibody binding interactions and neutralizing properties (20, 21). As preliminary MPXV assays showed variability and non-uniform plaque clusters (Fig S2), we introduced a sonication step. The sonication conditions of VACV and MPXV stocks used in our study were determined empirically by sonicating at increasing levels of intensity using a cup horn sonicator. While plaque titrations of MPXV without sonication produced visible clusters of plaques which prevented accurate titer estimation, even low levels of sonication treatment resulted in well separated MPXV plaques and significantly increased titers (p = 0.0166, Fig S2). We chose the third intensity setting “S3” for subsequent use as it was the lowest setting that provided significantly increased plaque numbers for both viruses (p = 0.0185, Fig S2). Virus was sonicated according to this procedure at the start of each PRNT.

We also considered the duration of virus incubation with sera prior to infection, as some PRNT studies of MPXV and VACV neutralization have extended the virus:sera incubation to overnight rather than one hour at 37°C (22, 23). We found that the extended incubation time was suboptimal despite producing increased PRNT_50_ titers because infectivity of the viruses also decreased independently of neutralizing antibodies with the extended adsorption time. MPXV demonstrated a 43.2% reduction in mean working dilution (p <0.00001) while VACV demonstrated a 20.9% reduction (p = 0.00121) (Fig S3B). This decreased infectivity suggested virus instability during the extended incubation time, which was greater for MPXV than VACV (Fig S3B).

Neutralizing antibody responses were measured for both MPXV and VACV by PRNT (Fig 2). The donor with prior smallpox vaccination had higher levels of neutralization than the naïve donors (Fig 2A-B) and is the only individual that produced a positive PRNT_90_. (Fig 2). Due to the difference in vaccination history, data points from this individual are shown in plots but were excluded from the overall mean PRNT titer calculations.

**Figure 2.**
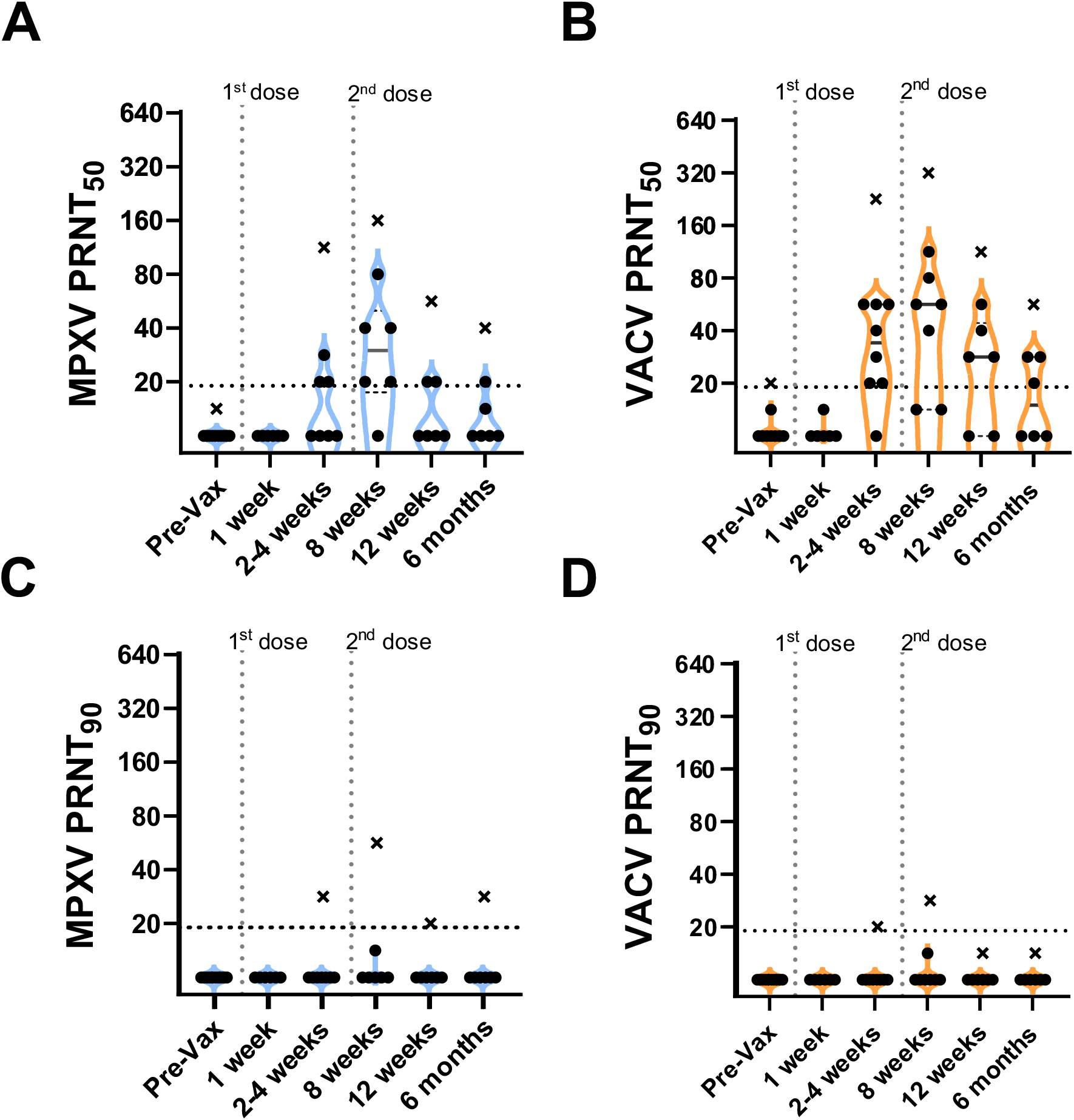
PRNT titers for participants vaccinated with JYNNEOS up to six months prior. Sera from donors vaccinated with two doses of JYNNEOS approximately 28 days apart were tested by PRNT. Assays were performed with sonicated virus and a one hour virus:sera incubation. A) MPXV PRNT_50_ and B) VACV PRNT_50_ C) MPXV PRNT_90_ and D) VACV PRNT_90_. Participants with no known vaccinia exposure (black circles) are used for mean calculations. Data from a single donor with prior smallpox vaccination (black Xs) are plotted separately and excluded from mean calculations. Each datapoint represents the geometric mean titer (GMT) of two independent experiments performed in duplicate. The vertical dotted lines represent the timing of the vaccine doses and the limits of detection (LOD) are expressed by horizontal dotted lines.

In all previously naïve individuals tested, the neutralizing antibody responses toward MPXV peaked at an average geometric mean PRNT_50_ titer (GMT) of 1:35 approximately one month following the 2^nd^ dose of JYNNEOS and quickly waned to below the 1:20 limit of detection (LOD) (Fig 2). Comparatively, neutralization of VACV was better than MPXV following just one dose of vaccine and was more robust with a peak GMT PRNT_50_ of 1:61 at eight weeks post initial dose (Fig 2-3). Neutralization of either virus waned similarly over time post-vaccination (Fig 2). One individual mounted no detectable neutralization response to MPXV (Fig. 2A). PRNT_50_ titers of most individuals to MPXV and VACV were below the limit of detection for both viruses by 12 weeks post initial vaccine dose (Fig 2A-B). At 12 months, post-vaccination sera from previously naïve individuals retained some reactivity to VACV with a PRNT_50_ GMT of 1:23, but neutralization of MPXV was at or below the PRNT_50_ LOD with a GMT of 1:12 (Fig 3). No naïve donors produced a detectable PRNT_90_ titer of at least 1:20 to either MPXV or VACV at any time point.

**Figure 3.**
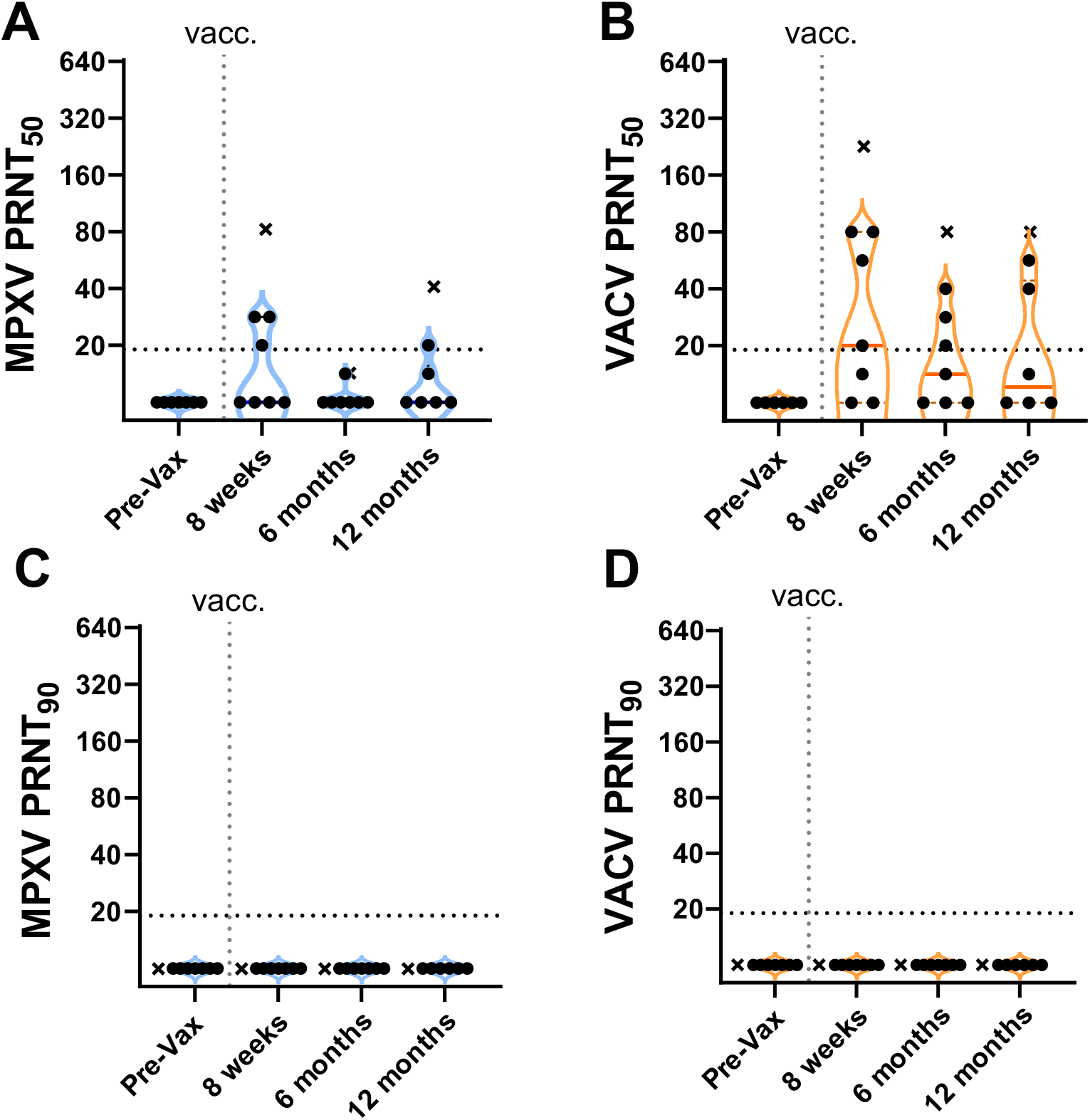
Longitudinal neutralizing antibody response by JYNNEOS vaccination extending to 12 months. Sera from donors vaccinated with two doses of JYNNEOS approximately 28 days apart were tested by PRNT. Assays were performed with sonicated virus and a one hour virus:sera incubation. A) MPXV PRNT_50_ and B) VACV PRNT_50_ C) MPXV PRNT_90_ and D) VACV PRNT_90_. Data from a single donor with prior smallpox vaccination are plotted separately (Xs). Data from participants with no known vaccinia exposure were used for mean calculations (black circles). Each datapoint represents the geometric mean titer (GMT) of two independent experiments, each performed in duplicate. Limit of detection (LOD) is expressed by horizontal dotted line.

## Discussion

The low levels of MPXV-neutralizing activity induced by JYNNEOS vaccination observed in this study are consistent with results of other recent studies, some of which have raised concerns over the efficacy and durability of MVA-BN vaccines in preventing mpox disease and spread (24, 25). We also found that neutralization titers can be impacted by assay conditions, which should be considered when comparing neutralizing activity levels from different studies. Empirical testing of MPXV PRNT assay conditions revealed that sonication can improve plaque quality and assay reliability, warranting its inclusion as part of a standardized protocol. In contrast, MPXV stability concerns argue against extending the pre-infection incubation period from one hour to overnight despite some increased sensitivity.

The donor who received prior ACAM2000 vaccination produced a greater IgG response and higher neutralization titers than naïve donors. However, the extent to which this individual’s neutralizing antibody response to MPXV was impacted by intrinsic differences between ACAM2000 and JYNNEOS, such as replication competence, versus the boosting of a memory response by additional vaccine doses is unclear. Other studies suggest that neutralizing antibody responses to MPXV can be enhanced by either a third MVA booster dose following the initial series or MVA vaccination after a first-generation smallpox vaccination (24, 26, 27). Both immunization strategies produced elevated neutralizing antibody levels to VACV that were stable when measured out to six months (28). MPXV neutralizing antibody levels can likewise be enhanced by both strategies, albeit to a lesser degree. One important caveat is that currently available studies measure MPXV antibody levels only shortly after boosting and thus do not address durability of the effect of prior vaccination history (24, 26).

There is evidence that first generation smallpox vaccines can produce a degree of a long-lasting humoral response to MPXV (24, 29, 30). However, long term studies on the durability of the MPXV-neutralizing antibody response derived from the second-generation vaccine ACAM2000 were not evaluated when the FDA established non-inferiority of JYNNEOS to ACAM2000 (31-33). While ACAM2000 is a replicating vaccinia strain derived from the live Dryvax vaccine, MVA-BN is a vaccinia strain which contains mutations and deletions that restrict its replication in human cells. Due to these differences, both the durability and the specificity of immune response elicited by MVA-BN may bear less similarity to historical smallpox vaccination than expected and should be evaluated further. For example, IgG levels induced by JYNNEOS vaccination exhibited a temporal response in naive vaccinees, which mirrored the waning neutralizing antibody response observed in this study and showed differential reactivity between VACV and MPXV antigens. Further testing is also needed to establish which antigens are important in eliciting neutralizing antibodies to MPXV and determine whether JYNNEOS preferentially elicits antibody responses to intracellular mature virions versus antigens associated with the extracellular envelope.

The possibility of a more efficacious cellular immune response to MPXV also warrants consideration. Cohn et al found that JYNNEOS vaccination led to an increase in CD4 and CD8 T cells that could recognize and respond to Orthopoxvirus specific antigens [14]. These CD4 and CD8 T cell responses from the JYNNEOS 2-dose recipients were similar to those of MPXV-convalescent donors. Cytokine responses were also comparable in the vaccinated versus convalescent groups. However, the protectiveness of these cellular responses and their longevity requires further study, as samples from vaccinated and convalescent donors were collected up to approximately four months post exposure. A comparative challenge study of MVA-BN and ACAM2000 immunized rhesus macaques also found that both vaccines produced similar T cell responses to VACV lysate (34).

Mpox remains a threat to public health, as transmission is an ongoing issue. While the 2022 global outbreak was driven by strains of MPXV Clade IIb, the more virulent Clade I MPXV strains have recently demonstrated the capacity for sexual transmission within endemic regions such as the Democratic Republic of Congo (35, 36). Documented cases of vaccine breakthrough infections and re-infection further underline the continuing need for effective means of preventing mpox transmission (37). Careful examination of current vaccination strategies and responses is urgently needed to ensure the long-term mitigation of mpox in endemic areas and prevention of ongoing global spread.

## Supporting information

manuscript text and figures

## Data Availability

All data produced in the present work are contained in the manuscript or supplemental materials.

## Acknowledgements

We gratefully acknowledge BEI Resources as a source for virus and the donors who generously contributed material. H. Dillenbeck, S. Griesemer and the Wadsworth Center Tissue Culture and Media Core provided outstanding technical assistance that greatly facilitated the work.

## About the Author

Dr. Phipps is a scientist in the Severe Respiratory Pathogen Laboratory of the Wadsworth Center, New York State Department of Health, Albany. Her research focuses on antibody responses to emerging viral pathogens and relevant vaccines

## References

1. Thornhill JP, Gandhi M, Orkin C. Mpox: The Reemergence of an Old Disease and Inequities. Annu Rev Med. 2023.

2. Gilchuk I, Gilchuk P, Sapparapu G, Lampley R, Singh V, Kose N, et al. Cross-Neutralizing and Protective Human Antibody Specificities to Poxvirus Infections. Cell. 2016;167(3):684-94.e9.

3. Hammarlund E, Lewis MW, Carter SV, Amanna I, Hansen SG, Strelow LI, et al. Multiple diagnostic techniques identify previously vaccinated individuals with protective immunity against monkeypox. Nat Med. 2005;11(9):1005–11.

4. Kennedy RB, Ovsyannikova I, Poland GA. Smallpox vaccines for biodefense. Vaccine. 2009;27 Suppl 4:D73–9.

5. FDA approves first live, non-replicating vaccine to prevent smallpox and monkeypox [press release]. fda.gov: U.S. Food and Drug Administration, 9/24/2019 2019.

6. EMA recommends approval of Imvanex for the prevention of monkeypox disease [press release]. European Medicines Agency, 7/22/2022 2022.

7. Dalton AF, Diallo AO, Chard AN, Moulia DL, Deputy NP, Fothergill A, et al. Estimated Effectiveness of JYNNEOS Vaccine in Preventing Mpox: A Multijurisdictional Case-Control Study - United States, August 19, 2022-March 31, 2023. MMWR Morb Mortal Wkly Rep. 2023;72(20):553–8.

8. Deputy NP, Deckert J, Chard AN, Sandberg N, Moulia DL, Barkley E, et al. Vaccine Effectiveness of JYNNEOS against Mpox Disease in the United States. N Engl J Med. 2023;388(26):2434–43.

9. Guagliardo SAJ, Kracalik I, Carter RJ, Braden C, Free R, Hamal M, et al. Monkeypox Virus Infections After 2 Preexposure Doses of JYNNEOS Vaccine - United States, May 2022-May 2024. MMWR Morb Mortal Wkly Rep. 2024;73(20):460–6.

10. Rosenberg ES, Dorabawila V, Hart-Malloy R, Anderson BJ, Miranda W, O’Donnell T, et al. Effectiveness of JYNNEOS Vaccine Against Diagnosed Mpox Infection - New York, 2022. MMWR Morb Mortal Wkly Rep. 2023;72(20):559–63.

11. Paredes MI, Ahmed N, Figgins M, Colizza V, Lemey P, McCrone JT, et al. Underdetected dispersal and extensive local transmission drove the 2022 mpox epidemic. Cell. 2024.

12. Hazra A, Zucker J, Bell E, Flores J, Gordon L, Mitjà O, et al. Mpox in people with past infection or a complete vaccination course: a global case series. Lancet Infect Dis. 2023.

13. Yates JL, Hunt DT, Kulas KE, Chave KJ, Styer L, Chakravarthi ST, et al. Development of a novel serological assay for the detection of mpox infection in vaccinated populations. J Med Virol. 2023;95(10):e29134.

14. Hooper JW, Thompson E, Wilhelmsen C, Zimmerman M, Ichou MA, Steffen SE, et al. Smallpox DNA vaccine protects nonhuman primates against lethal monkeypox. J Virol. 2004;78(9):4433–43.

15. Cohn H, Bloom N, Cai GY, Clark JJ, Tarke A, Bermúdez-González MC, et al. Mpox vaccine and infection-driven human immune signatures: an immunological analysis of an observational study. Lancet Infect Dis. 2023;23(11):1302–12.

16. Lee WT, Girardin RC, Dupuis AP, Kulas KE, Payne AF, Wong SJ, et al. Neutralizing Antibody Responses in COVID-19 Convalescent Sera. J Infect Dis. 2021;223(1):47–55.

17. Beaty BJ CC, Shope RE. Arboviruses. In: Lennette EH LD, Lennette ET, editor. Diagnostic Procedures for Viral, Rickettsial, and Chlamydial Infections. Washington, DC: American Public Health Association; 1995. p. 189–212.

18. Galasso GJ, Sharp DG. Virus particle aggregation and the plaque-forming unit. J Immunol. 1962;88:339–47.

19. Kotwal GJ, Abrahams MR. Growing poxviruses and determining virus titer. Methods Mol Biol. 2004;269:101–12.

20. Burton DR. Antiviral neutralizing antibodies: from in vitro to in vivo activity. Nat Rev Immunol. 2023:1-15.

21. Pradhan S, Varsani A, Leff C, Swanson CJ, Hariadi RF. Viral Aggregation: The Knowns and Unknowns. Viruses. 2022;14(2).

22. Newman FK, Frey SE, Blevins TP, Mandava M, Bonifacio A, Jr., Yan L, Belshe RB. Improved assay to detect neutralizing antibody following vaccination with diluted or undiluted vaccinia (Dryvax) vaccine. J Clin Microbiol. 2003;41(7):3154–7.

23. Priyamvada L, Carson WC, Ortega E, Navarra T, Tran S, Smith TG, et al. Serological responses to the MVA-based JYNNEOS monkeypox vaccine in a cohort of participants from the Democratic Republic of Congo. Vaccine. 2022;40(50):7321–7.

24. Zaeck LM, Lamers MM, Verstrepen BE, Bestebroer TM, van Royen ME, Götz H, et al. Low levels of monkeypox virus-neutralizing antibodies after MVA-BN vaccination in healthy individuals. Nat Med. 2023;29(1):270–8.

25. Moschetta N, Raccagni AR, Bianchi M, Diotallevi S, Lolatto R, Candela C, et al. Mpox neutralising antibodies at 6 months from mpox infection or MVA-BN vaccination: a comparative analysis. The Lancet Infectious Diseases. 2023;23(11):e455–e6.

26. Hubert M, Guivel-Benhassine F, Bruel T, Porrot F, Planas D, Vanhomwegen J, et al. Complement-dependent mpox-virus-neutralizing antibodies in infected and vaccinated individuals. Cell Host Microbe. 2023;31(6):937–48 e4.

27. Shamier MCVL, L.P.M. Verstrepen, B.E. Wijnans, K. Ahkiyate, N. Gotz, H.M., De Vries R.D., Van Gorp, E.C.M. Koopmans M.P.G., Goeijenbier S, Geurtsvankessel CH, Zaeck, LM. Mpox-specific antibodies wane within one year after MVA-BN vaccination. 34th European Congress of Clinical Microbiology and Infectious Diseases; 2024.

28. Ilchmann H, Samy N, Reichhardt D, Schmidt D, Powell JD, Meyer TPH, et al. One- and Two-Dose Vaccinations With Modified Vaccinia Ankara-Bavarian Nordic Induce Durable B-Cell Memory Responses Comparable to Replicating Smallpox Vaccines. J Infect Dis. 2023;227(10):1203–13.

29. Li E, Guo X, Hong D, Gong Q, Xie W, Li T, et al. Duration of humoral immunity from smallpox vaccination and its cross-reaction with Mpox virus. Signal Transduct Target Ther. 2023;8(1):350.

30. Asquith W, Hueston L, Dwyer D, Kok J, Ko D, Fennel M, et al. Characterizing the acute antibody response of monkeypox and MVA-BN vaccine following an Australian outbreak. J Med Virol. 2024;96(1):e29407.

31. ACAM2000: smallpox vaccine (briefing document): Vaccines and Related Biological Products Advisory Committee; April 18, 2007 [Available from: https://wayback.archive-it.org/7993/20170405044400/ https://www.fda.gov/ohrms/dockets/ac/07/briefing/2007-4292B2-02.pdf.

32. Pittman PR, Hahn M, Lee HS, Koca C, Samy N, Schmidt D, et al. Phase 3 Efficacy Trial of Modified Vaccinia Ankara as a Vaccine against Smallpox. N Engl J Med. 2019;381(20):1897–908.

33. JYNNEOS (Smallpox and Monkeypox Vaccine, Live, Nonreplicating) [package insert], Hellerup, Denmark: Bavarian Nordic; 2023.

34. Jacob-Dolan C, Ty D, Hope D, McMahan K, Liu J, Powers OC, et al. Comparison of the immunogenicity and protective efficacy of ACAM2000, MVA, and vectored subunit vaccines for Mpox in rhesus macaques. Sci Transl Med. 2024;16(740):eadl4317.

35. Sun Y, Nie W, Tian D, Ye Q. Human monkeypox virus: Epidemiologic review and research progress in diagnosis and treatment. Journal of Clinical Virology. 2024;171:105662.

36. Kibungu E, Vakaniaki E, Kinganda-Lusamaki E, Kalonji-Mukendi T, Pukuta E, Hoff N, et al. Clade I–Associated Mpox Cases Associated with Sexual Contact, the Democratic Republic of the Congo. Emerging Infectious Disease journal. 2024;30(1):172.

37. Hazra A, Zucker J, Bell E, Flores J, Gordon L, Mitjà O, et al. Mpox in people with past infection or a complete vaccination course: a global case series. The Lancet Infectious Diseases. 2024;24(1):57–64.

