## Supplementary material for "JYNNEOS vaccination induced short-lived neutralizing antibody responses to monkeypox virus in naïve individuals": manuscript text and figures

### Supplemental Table and Figures

**Supplemental Table S1. Impact of sonication on number of plaque forming units**

| | | **Monkeypox Virus, USA-2003** | **Vaccinia Virus, Western Reserve** |
|---|---|---|---|
| Treatment[a] | Sonicator power setting | Mean fold change of sonicated versus non-sonicated condition, range (low -high)[b] | Mean fold change of sonicated versus non-sonicated condition, range (low -high)[b] |
| S2 | 2 | 5.22, (3.1 - 7.68) | 4.72, (4.50 – 5.16) |
| S3 | 3 | 9.59, (5.28 – 15.19) | 4.81, (3.64 – 6.93) |
| S4 | 4 | 9.82, (8.44 – 11.81) | 5.74, (4.57 – 7.45) |
| S5 | 5 | 8.19, (5.28 – 11.72) | 4.13, (2.64 – 6.05) |

[a]Monkeypox virus (MPXV) and vaccinia virus (VACV) virus aliquots were sonicated at intensity settings 2, 3, 4, or 5 and subsequently titrated by plaque assay.

[b]The fold change of each biological replicate was determined by dividing the virus titer of sonication treated aliquots by the non-sonicated aliquot titer. The mean fold change is the result of three biological replicates.

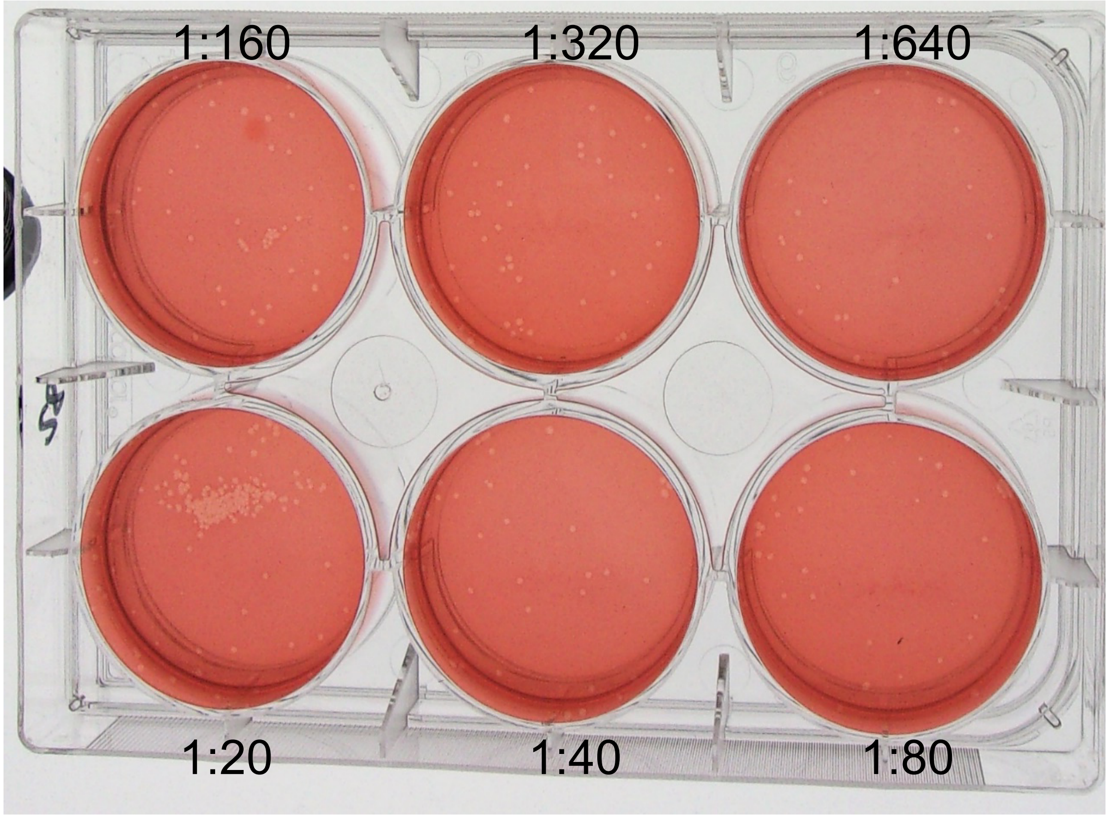


**Figure S1. Plaque clusters in MPXV titration well.**

Plaques shown for multiple serum dilutions (1:20 through 1:640) in a PRNT of MPXV performed without sonicating virus before inoculation were non-uniform, producing a large plaque cluster in bottom left (1:20) well and small clusters in top left (1:160) well. Image brightness, contrast, and saturation were optimized for visualization.

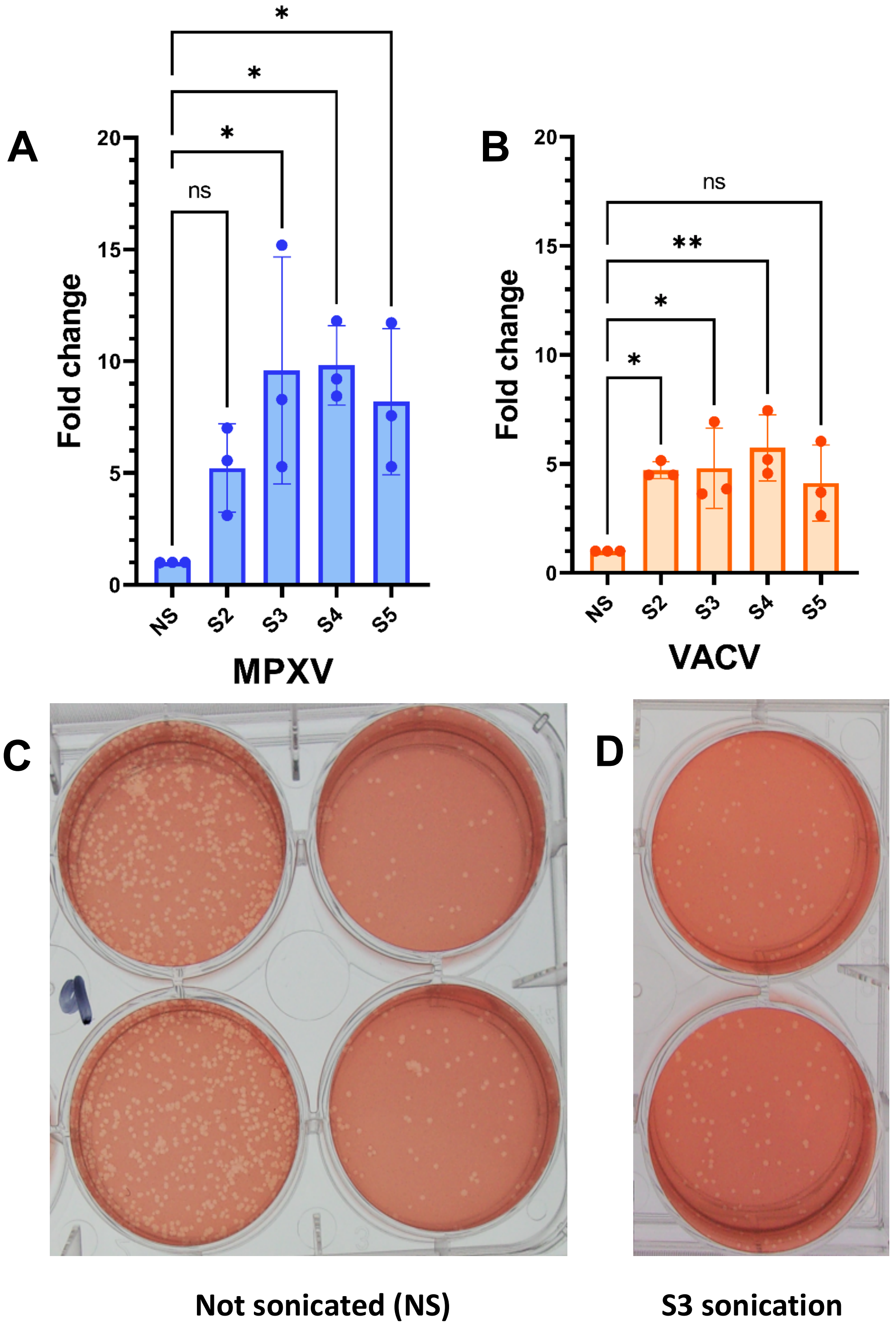


**Figure S2. Optimization of MPXV and VACV sonication.**

Fold change in virus titer normalized to non-sonicated virus titer for A) MPXV and B) VACV. The mean represents the average of three biological replicates as in Table S1. C) Plaque assays of MPXV showing plaque morphology when not sonicated and D) with sonication treatment at setting 3. One-way analysis of variance (ANOVA) with multiple comparisons was used to evaluate significance relative to non-sonicated condition. *, P <0.05, ** P, <0.005. Image brightness, contrast, and saturation were optimized for visualization.

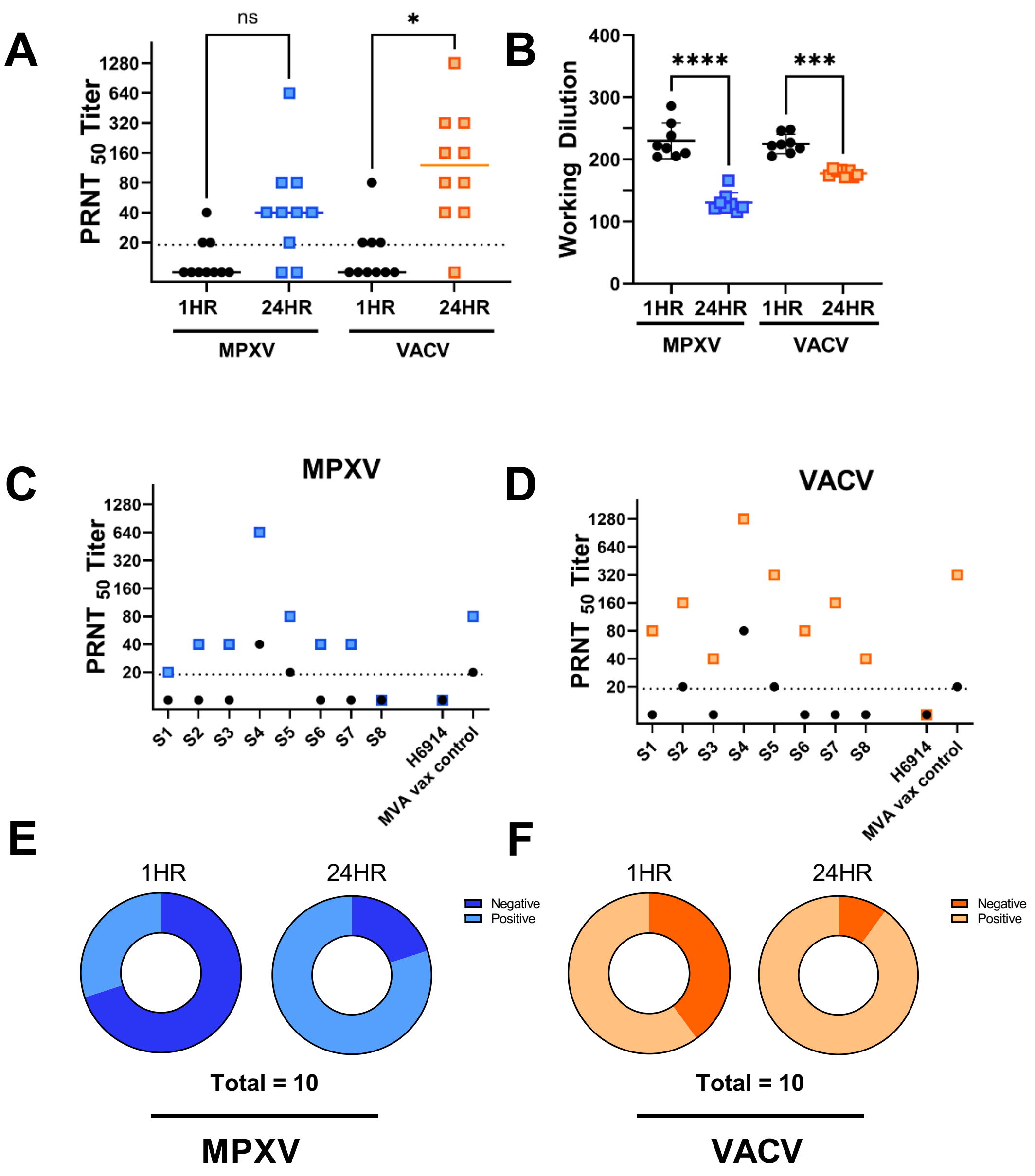


**Figure S3. Evaluation of extended virus:sera adsorption period.**

Eight independent serum samples of various $PRNT_{50}$ titers were selected and positive and negative controls were tested in paralell by performing a PRNT with a virus:sera incubation step of either one or twenty four hours at 37°C. The negative control consisted of pooled serum product #H61914-20ML Human serum type AB (H6914). The "MVA vax control" used was a donor sample of which the PRNT range had been previously determined through at least three independent assays. $PRNT_{50}$ titers A) Combined $PRNT_{50}$ values for MPXV or VACV B) Working dilution was determined by back titration of virus inoculum in media alone following one and twenty-four hours incubation alongside virus:sera samples. After twenty-four hours incubation, MPXV and VACV inoculum titers were reduced by 43.2 % and 20.9% respectively as compared to the titer of the same virus preparation when incubated for one hour. Individual data from the summary in A) are shown in C) for MPXV and D) for VACV. $PRNT_{50}$ titers above 1:20 are considered seropositive by the limit of detection of our assay and are displayed in lighter colors of E) for MPXV and F) for VACV. One-way analysis of variance (ANOVA) with multiple comparisons was used to compare 1HR and 24HR conditions. *, P <0.05, *, P <0.05, ** P, <0.005, ***, P <0.0005, ****, P < 0.00005
